# Glycemic patterns from continuous glucose monitoring devices among women with regular and irregular menstrual cycles

**DOI:** 10.64898/2026.10.16.26365453

**Authors:** Zifan Wang, Gowtham Asokan, Boya Zhang, Sophia Werdin, Paola Bojorquez-Ramirez, Sanaa H. Suharwardy, Jukka-Pekka Onnela, Michelle A. Williams, Brent A. Coull, Shruthi Mahalingaiah

## Abstract

Polyendocrine metabolic ovarian syndrome (PMOS) and early-life irregular cycles (ELIC) carry higher metabolic risks, but real-world glycemic patterns remain inadequately characterized. We analyzed 247,316 continuous glucose monitoring (CGM) days from 716 consented and enrolled participants (11/2019-08/2026) in the Apple Women’s Health Study, using cluster-weighted generalized estimating equations to evaluate associations between PMOS, ELIC, and glycemic metrics. Same-day wearable-assessed physical activity, digital dietary logs, and medication health records were assessed as effect modifiers. Compared to participants without PMOS and with regular cycles, those with PMOS had higher mean glucose (6.81 mg/dL; 95% CI: −0.70, 14.72) and %observations above target range (3.77%; 95% CI: 0.04, 8.14). Estimates were smaller-in-magnitude for participants without PMOS but with ELIC, and larger for PMOS with androgen excess. Meeting activity goals, consuming protein-/fiber-rich diets, or using metformin/GLP-1 receptor agonists attenuated these associations. PMOS/ELIC were associated with unfavorable glycemic profiles, suggesting opportunities for risk identification and modification.

## Introduction

The menstrual cycle is increasingly recognized as a vital sign from menarche through menopause^1^. Polycystic ovary syndrome (PCOS) is a common endocrine condition affecting around 5–18% of reproductive-aged women, characterized by irregular menstrual cycles and hyperandrogenism^2^. In May 2026, PCOS was renamed as polyendocrine metabolic ovarian syndrome (PMOS), to reflect its metabolic complexity and known associations with insulin resistance, prediabetes, and type 2 diabetes^3–5^; we use the terminology PCOS/PMOS hereinafter for continuity with the existing literature and the survey instrument wording used in our work. Not all individuals with irregular menstrual cycles have or receive a diagnosis of PCOS/PMOS, due to underdiagnosis or other endocrinopathies^6^. Prolonged time from menarche to cycle regularity is an early marker for later-life menstrual irregularity, and is independently associated with increased prevalence of prediabetes, type 2 and type 1 diabetes, and gestational diabetes, even among those without a PCOS/PMOS diagnosis^4,7^. However, real-world glycemic patterns beyond a diagnosis indicator, or the role of potential modifiable lifestyle factors in these glycemic patterns, have not been well characterized among individuals with PCOS/PMOS and/or early-life menstrual irregularity.

Continuous glucose monitoring (CGM) devices provide more detailed, real-time glycemic patterns and glucose metrics (such as time spent in range and glucose variability) beyond traditional measures like hemoglobin A1c (HbA1c) or fingerstick glucose testing^8^. In addition to becoming a standard of care for type 1 diabetes management and being recommended for type 2 diabetes self-management^8,9^, in recent years, CGM use has been expanding to populations without diabetes for metabolic risk assessment, performance optimization (e.g., for athletes), and general health and wellness monitoring^10^. This expansion may accelerate in the United States, with the U.S. Food and Drug Administration’s clearance of the first, over-the-counter CGM in March 2024^11^.

Limited studies have evaluated CGM-derived glycemic patterns in relation to PCOS/PMOS, and most were pilot or feasibility studies. One study demonstrated feasibility of CGM use (42-day wear) among 30 participants with PCOS/PMOS, and found higher daytime glucose in those with hyperandrogenism compared with those without^12^, consistent with another study (n=53, 48-hour wear) showing higher mean glucose in those with hyperandrogenism^13^. One study (14-day CGM wear among 36 women with PCOS/PMOS) observed lower glucose variability among those with insulin resistance^14^. Lastly, a US-based study (serial 24-hour CGM) comparing pregnant women with and without PCOS/PMOS (n=34) found subtle differences in postprandial glucose^15^. A few other studies among women with type 1 diabetes (sample sizes: 4 to 124) examined CGM-derived glycemic measures by menstrual cycle phases (follicular vs luteal) among women with type 1 diabetes^16^, and one study among 49 non-diabetic Canadian participants found biphasic patterns of glucose across the menstrual cycle^17^, but none evaluated how PCOS/PMOS or irregular cycles may impact glucose patterns. Existing studies were typically conducted in controlled settings with specified CGM wear time, and recruited small, convenience-sampled groups from clinics or specific locations, often limiting generalizability or statistical power to fully investigate associations. To our knowledge, no population-based study has comprehensively evaluated the associations between PCOS/PMOS and early-life menstrual irregularity and glycemic patterns from real-world CGM use data, and evidence regarding potential lifestyle modifications to improve glycemic control in these populations remains sparse.

In this study, we analyzed 247,316 days of CGM data among 716 participants in a US-based digital cohort to examine daily glycemic outcomes across three exposure groups: a regular cycles group, consisting of those without diagnosed PCOS/PMOS and reporting early-life regular cycles; a irregular cycles group, consisting of those without diagnosed PCOS/PMOS but reporting early-life irregular cycles; and a PCOS/PMOS group, consisting of those reporting a PCOS/PMOS diagnosis. As secondary analyses, we evaluated wearable-derived physical activity, self-logged dietary patterns, and medication use from health records as potential effect modifiers for the associations between PCOS/PMOS or early-life menstrual irregularity and glycemic outcomes. A conceptual model of the primary and secondary research questions and potential clinical and public health implications is shown in Figure 1, with participant flowchart shown in Supplemental Figure S1.

**Figure 1.**
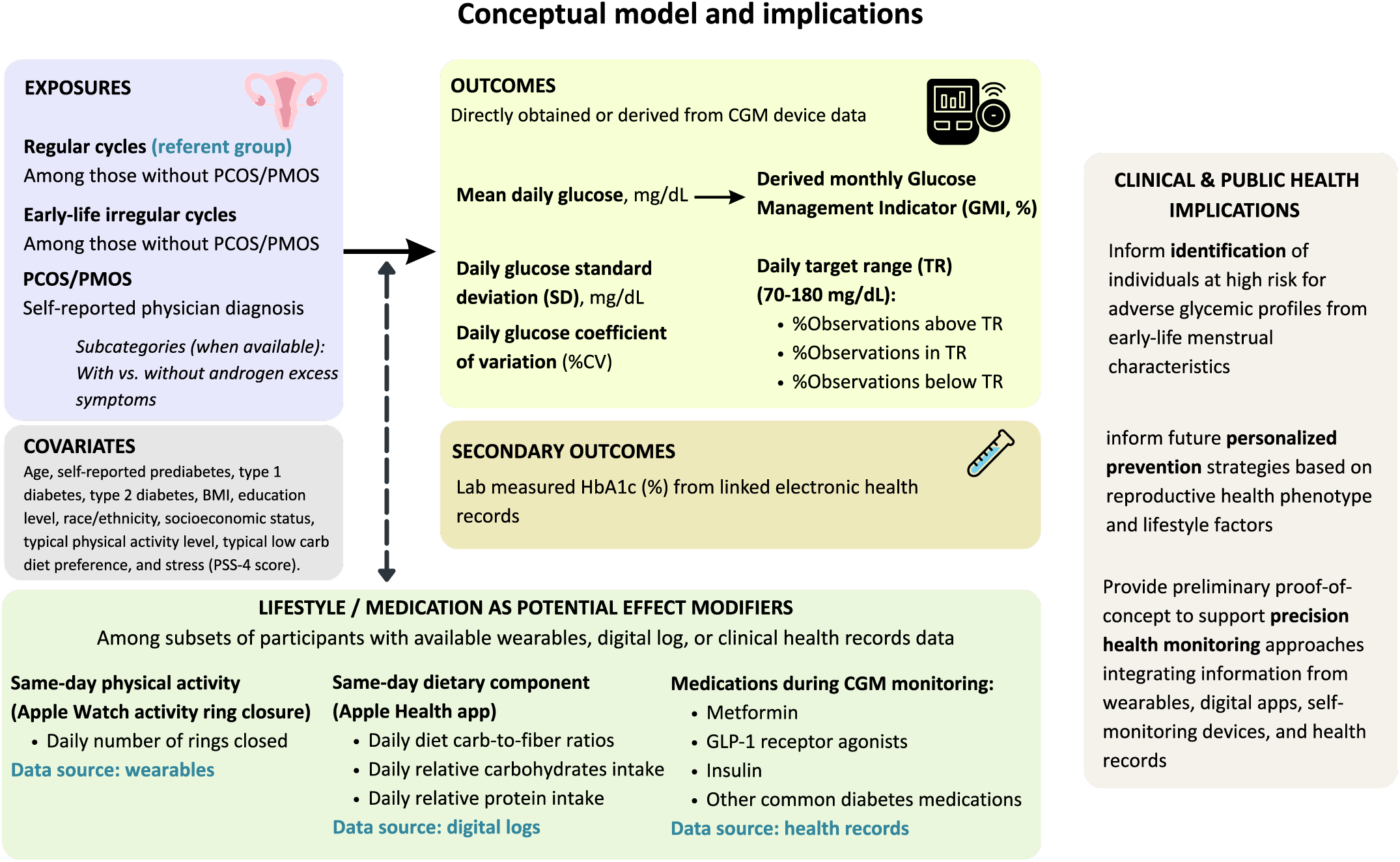
Conceptual model and implications. Abbreviations: CGM, continuous glucose monitoring; GLP-1 RA, glucagon-like peptide-1 receptor agonist; PCOS, polycystic ovary syndrome [in May 2026, PCOS was renamed as polyendocrine metabolic ovarian syndrome (PMOS)]. Early-life regular cycles group: those without self-reported PCOS/PMOS diagnosis, and reported spontaneously establishing regular cycles within 4 years after menarche; Early-life irregular cycles group: those without self-reported PCOS/PMOS diagnosis, and reported reaching cycle regularity in 5+ years after menarche, their cycles are still irregular, or used hormones to achieve regularity.

## Results

### Participant characteristics

Participant characteristics are shown in Table 1. Among the 716 participants who provided consent and opted in to share relevant data with the Study, the referent group (regular cycles) were of older age (mean: 41.2 years) compared to the irregular cycles (mean: 36.4 years) or PCOS/PMOS group (mean: 37.4 years). Around 74-79% self-identified as non-Hispanic White. The three groups substantially differed by socioeconomic status (SES), education, body mass index (BMI), and preexisting prediabetes/diabetes status, with the referent group reporting higher SES and college or graduate degree, and the PCOS/PMOS group having higher prevalence of obesity, prediabetes, and type 2 diabetes. For summary statistics of glycemic outcomes, the irregular cycles or PCOS/PMOS groups had higher mean glucose and %observations above target range compared to the referent group. Other supporting data in Table 1 suggests that the PCOS/PMOS group had lower daily prevalence of meeting all physical activity goals or consuming fiber-rich diet, while they reported higher usage of metformin or glucagon-like peptide-1 receptor agonists (GLP-1 RAs).

**Table 1.** Characteristics of the 716 participants by exposure group.

| Participant characteristics<sup>a</sup> | Mutually exclusive exposure groups |  |  |
| --- | --- | --- | --- |
|  | No PCOS/PMOS |  | PCOS/PMOS<br>(N = 167) |
|  | Regular cycles<br>(N = 412) | Irregular cycles<br>(N = 137) |  |
| **Baseline covariate distributions** |  |  |  |
| **Age in years:** |  |  |  |
| Median (IQR) | 41 (32 – 50) | 36 (28 – 44) | 37 (31 – 43) |
| Mean ± SD | 41.2 ± 12.7 | 36.4 ± 10.4 | 37.4 ± 9.2 |
| **Race/ethnicity, n (%)** |  |  |  |
| Non-Hispanic White | 309 (75.0) | 108 (78.8) | 123 (73.7) |
| Non-Hispanic Black | 25 (6.1) | 15 (11.9)<sup>b</sup> | 20 (12.0)<sup>b</sup> |
| Hispanic | 23 (5.6) |  |  |
| Multiple, Asian, American Indian or Alaska Native,<br>Middle Eastern or North African, Native Hawaiian or<br>Pacific Islander, or other | 54 (13.1) | 13 (9.5) | 24 (14.4) |
| **Subjective SES scale, n (%)** |  |  |  |
| Low (scale: 0–3) | 105 (25.5) | 48 (35.0) | 63 (37.7) |
| Medium (scale: 4–5) | 164 (39.8) | 56 (40.9) | 71 (42.5) |
| High (scale: 6–9) | 141 (34.2) | 32 (23.4) | 33 (19.8) |
| **Education, n (%)** |  |  |  |
| ≤High school | 42 (10.2) | 25 (18.2) | 21 (12.6) |
| Some college or tech school | 126 (30.6) | 54 (39.4) | 76 (45.5) |
| College degree | 118 (28.6) | 36 (26.3) | 34 (20.4) |
| Graduate school | 126 (30.6) | 22 (16.1) | 36 (21.6) |
| **BMI categories, n (%)** |  |  |  |
| <25 kg/m<sup>2</sup> | 109 (26.5) | 35 (25.5) | 17 (10.2) |
| 25–29.9 kg/m<sup>2</sup> | 97 (23.5) | 35 (25.5) | 26 (15.6) |
| ≥30 kg/m<sup>2</sup> | 201 (48.8) | 64 (46.7) | 122 (73.1) |
| **Diabetes status (self-reported physician diagnosis):** |  |  |  |
| Prediabetes, n (%)<sup>c</sup> | 53 (12.9) | 17 (12.4) | 42 (25.1) |
| Type 1 diabetes, n (%)<sup>c</sup> | 156 (37.9) | 59 (43.1) | 39 (23.4) |
| Type 2 diabetes, n (%)<sup>c</sup> | 113 (27.4) | 42 (30.7) | 85 (50.9) |
| No prediabetes/diabetes, n (%)<sup>c</sup> | 122 (29.6) | 24 (17.5) | 27 (16.2) |
| Self-reported typical **exercise minutes >150**<br>**minutes/week**, n (%) | 97 (23.5) | 26 (19.0) | 21 (12.6) |
| Self-reported **typical low carbohydrate diet**, n (%) | 112 (27.2) | 42 (30.7) | 64 (38.3) |
| **Perceived Stress Scale 4 (PSS-4)**, mean ± SD | 6.2 ± 3.6 | 7.5 ± 3.7 | 7.3 ± 3.4 |
| **Hormone ever use** (for managing cycles,<br>contraception, infertility treatment, menopause, or<br>other reasons), n (%) | 316 (76.7) | 111 (81.0) | 149 (89.2) |
| **CGM data summary statistics** |  |  |  |
| **Data density:** |  |  |  |
| Number of days per person, median (IQR) | 195 (52 – 507) | 242 (66 – 606) | 209 (57 – 487) |
| Number of glucose readings per day, median (IQR) | 288 (288 – 288) | 288 (288 – 288) | 288 (288 – 288) |
| **Summary of within-individual glycemic measures:** |  |  |  |
| Mean glucose (mg/dL), mean ± SD | 147.0 ± 40.7 | 159.1 ± 48.5 | 155.6 ± 48.6 |
| Mean percent of observed glucose readings in target<br>range (%), mean ± SD | 76.9 ± 22.1 | 70.8 ± 25.5 | 73.0 ± 25.6 |
| Mean percent of observed glucose readings above<br>target range (%), mean ± SD | 21.5 ± 22.2 | 27.9 ± 25.6 | 26.0 ± 26.0 |
| Mean percent of observed glucose readings below target range (%), mean ± SD | 1.5 ± 2.0 | 1.3 ± 1.9 | 1.1 ± 2.2 |
| Mean glucose standard deviation (SD) , mean ± SD | 34.7 ± 21.6 | 38.2 ± 24.3 | 34.4 ± 19.6 |
| Mean coefficient of variation (CV, %), mean ± SD | 22.5 ± 6.5 | 23.6 ± 8.3 | 21.1 ± 6.6 |
| **Effect modifiers evaluated** |  |  |  |
| **Number of same-day Apple Watch activity ring closed per day among a subset of 521 participants with eligible Apple Watch data, with 113,324 days during CGM monitoring, n (%) days:** |  |  |  |
| 0 activity rings closed | 8173 days<br>(12.1) | 3862 days<br>(15.5) | 3400 days<br>(16.1) |
| 1 activity ring closed | 17426 days<br>(25.9) | 6275 days<br>(25.2) | 6524 days<br>(30.9) |
| 2 activity rings closed | 22797 days<br>(33.9) | 8783 days<br>(35.2) | 7592 days<br>(36.0) |
| All 3 activity rings closed | 18913 days<br>(28.1) | 6008 days<br>(24.1) | 3571 days<br>(16.9) |
| **Same-day dietary macronutrient composition among a subset of 58 participants with digitally logged dietary data, with 2,565 days during CGM monitoring, n (%) days:** |  |  |  |
| Diet rich in fiber (carbohydrates-to-fiber ratio ≤ 10:1) | 724 days (70.4) | 659 days (60.6) | 233 days (51.8) |
| Diet with relatively low carbohydrates (<40% of total energy consumed) | 521 days (50.7) | 393 days (36.2) | 258 days (57.3) |
| Diet with relatively high/moderate protein (≥12% of total energy consumed) | 762 days (74.1) | 706 days (64.9) | 365 days (81.1) |
| **Medications use among a subset of 180 participants with shared electronic health records, with 65,885 days during CGM monitoring, n (%) days:** |  |  |  |
| Metformin used | 4669 days<br>(13.6) | 3290 days<br>(19.7) | 4381 days<br>(29.3) |
| GLP-1 RA used | 4741 days<br>(13.9) | 2873 days<br>(17.2) | 3212 days<br>(21.5) |
| Insulin used | 16848 days<br>(49.2) | 12293 days<br>(73.5) | 5662 days<br>(37.9) |
| Not any common diabetes medications used | 13409 days<br>(39.2) | 3545 days<br>(21.2) | 7185 days<br>(48.1) |
| **Cardiometabolic vital signs** |  |  |  |
| Within-individual average of HbA1c (%hemoglobin) among 139 participants with clinical HbA1c lab records within +/- 1 month around any of their CGM readings, mean ± SD | 6.8 ± 1.4 | 7.4 ± 1.6 | 7.4 ± 2.2 |
| Within-individual average of daily resting heart rate (bpm) among 567 participants with 117,931 same-day resting heart rate data from Apple Watch during CGM monitoring, mean ± SD | 70.6 ± 9.7 | 70.3 ± 9.1 | 74.5 ± 8.9 |

Abbreviations: EL: early-life; IQR, interquartile range; SD, standard; CGM, continuous glucose monitoring; GLP-1 RA, glucagon-like peptide-1 receptor agonists; PCOS, polycystic ovary syndrome. In May 2026, PCOS was renamed as polyendocrine metabolic ovarian syndrome (PMOS). We use PCOS throughout this manuscript because it was the term used when survey data were collected, prior to the May 2026 renaming.
<sup>a</sup> Numbers may not add up fully to the total N due to missingness (missingness between 0–1.4%).
<sup>b</sup> Categories with <10 participants aggregated to enhance data confidentiality.
<sup>c</sup> The numbers of participants reporting physician diagnosis of prediabetes, type 1 diabetes, type 2 diabetes, and no prediabetes/diabetes sum to more than the total number of participants, because the survey questions asked whether participants had *ever* received each diagnosis and allowed them to select all applicable responses. Specifically, 49 participants reported having ever received diagnoses of both prediabetes and type 2 diabetes, which is plausible when prediabetes preceded the later development of type 2 diabetes. Twelve participants either reported having received diagnoses of both type 1 and type 2 diabetes, or reported diagnoses of both prediabetes and type 1 diabetes (only 2.2% among all 543 participants with any prediabetes/diabetes); such overlaps are feasible, reflecting potential diagnostic reclassification, or the coexistence of type 1 diabetes with insulin resistance and other metabolic features of type 2 diabetes [American Diabetes Association Professional Practice Committee for Diabetes\*; 2. Diagnosis and Classification of Diabetes: Standards of Care in Diabetes—2026. *Diabetes Care* 1 January 2026; 49 (Supplement\_1): S27–S49].

### Associations between PCOS/PMOS, early-life cycle irregularity, and glycemic patterns

In covariate-adjusted cluster-weighted GEE models (estimates and 95% CIs shown in Figure 2), compared to the referent group: 1) participants with PCOS showed higher daily mean glucose [β (95% CI) = 6.81 (−0.70, 14.72) mg/dL], lower %observations in target range [β (95% CI) = −3.62 (−8.01, 0.04) %], higher %observations above target range [β (95% CI) = 3.77 (0.04, 8.14) %], and higher glucose management indicator (GMI) derived from CGM values [β (95% CI) = 0.20 (0.00, 0.42) %], consistently suggesting a trend towards higher glucose patterns; 2) participants with irregular cycles had higher daily mean glucose [β (95% CI) = 6.33 (−1.48, 14.40) mg/dL], lower %observations in target range [β (95% CI) = −3.08 (−7.11, 0.91) %], higher %observations above target range [β (95% CI) = 3.33 (−0.77, 7.41) %], and higher derived GMI [β (95% CI) = 0.15 (−0.01, 0.34) %], although the effect estimates’ magnitudes were smaller than that among the PCOS/PMOS group. No strong associations were observed with glucose variability (SD or %CV) for either group when adjusted for covariates (Supplemental Table S1).

**Figure 2.**
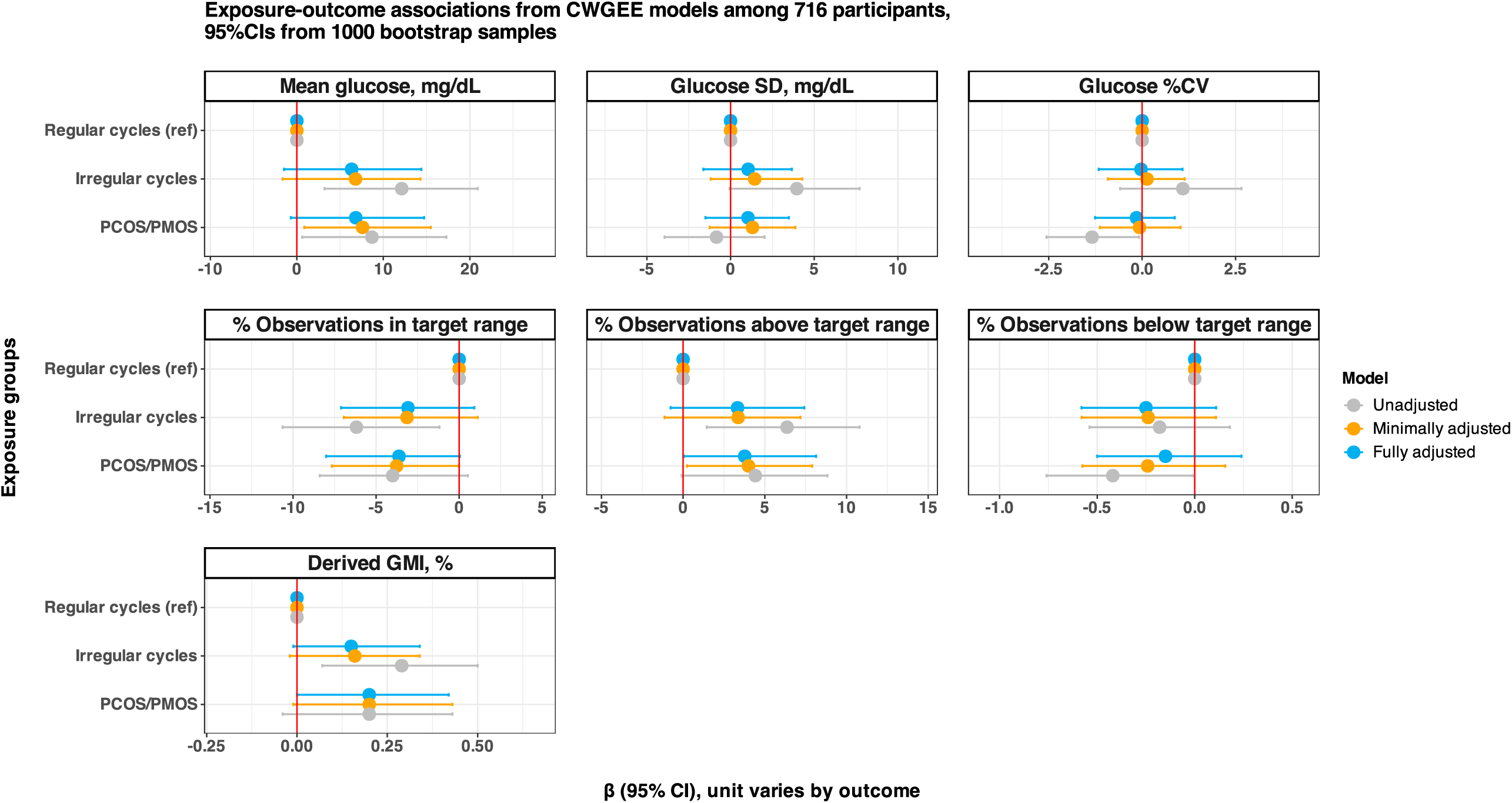
Effect estimates and bootstrap 95% CIs from CWGEE models. Abbreviations: CGM, continuous glucose monitoring; CI, confidence interval; CWGEE, cluster-weighted generalized estimating equations; GMI, glucose management indicator; PCOS, polycystic ovary syndrome [in May 2026, PCOS was renamed as polyendocrine metabolic ovarian syndrome (PMOS)]. Regular cycles group: those without self-reported PCOS diagnosis, and reported spontaneously establishing regular cycles within 4 years after menarche; Irregular cycles group: those without self-reported PCOS diagnosis, and reported reaching cycle regularity in 5+ years after menarche, their cycles are still irregular, or used hormones to achieve regularity. Red vertical line represents β = 0 as the referent group. Bias-corrected bootstrap confidence intervals were estimated from 1000 bootstrap samples. Minimally adjusted models adjusted for age, self-reported prediabetes, type 1 diabetes, and type 2 diabetes. Fully adjusted models adjusted for age, self-reported prediabetes, type 1 diabetes, type 2 diabetes, BMI, education level, race/ethnicity, socioeconomic status, typical physical activity level, typical low carb diet preference, and stress reflected by PSS-4 score. All covariates are self-reported at enrollment.

### PCOS/PMOS or early-life cycle irregularity and HbA1c values among subsets with electronic health records (EHRs)

Among subsets of 139 to 227 participants who linked and shared their electronic health records, unadjusted point estimates suggest participants with PCOS/PMOS or irregular cycles had 0.47 %hemoglobin (95% CI: 0.03, 0.89) higher mean HbA1c value from lab records before their CGM readings (i.e., long-term HbA1c preceding CGM), while this association attenuated to 0.17 (95% CI: −0.30, 0.66) after adjusting for covariates; participants with PCOS/PMOS or irregular cycles had 0.58 %hemoglobin (95% CI: 0.03, 1.22) higher mean HbA1c value from lab records measured around +/− 1 month around their CGM readings (i.e., concurrent short-term HbA1c values) in unadjusted CWGEE models, although this association attenuated to 0.36 (−0.22, 0.89) after covariate adjustment (Supplemental Table S2). Among the 139 participants with concurrent short-term EHR-based HbA1c values along with GMI derived from their CGM values, their EHR-based HbA1c values had acceptable agreement with their CGM-derived GMI, with a Spearman correlation coefficient of 0.84 (Supplemental Figure S2).

### Potential effect modifications by physical activity, diet, and medication use

When evaluating associations within subgroups defined by the potential effect modifiers, Figures 3-5 show key findings relevant to potential effect modification from stratified analyses.

**Figure 3.**
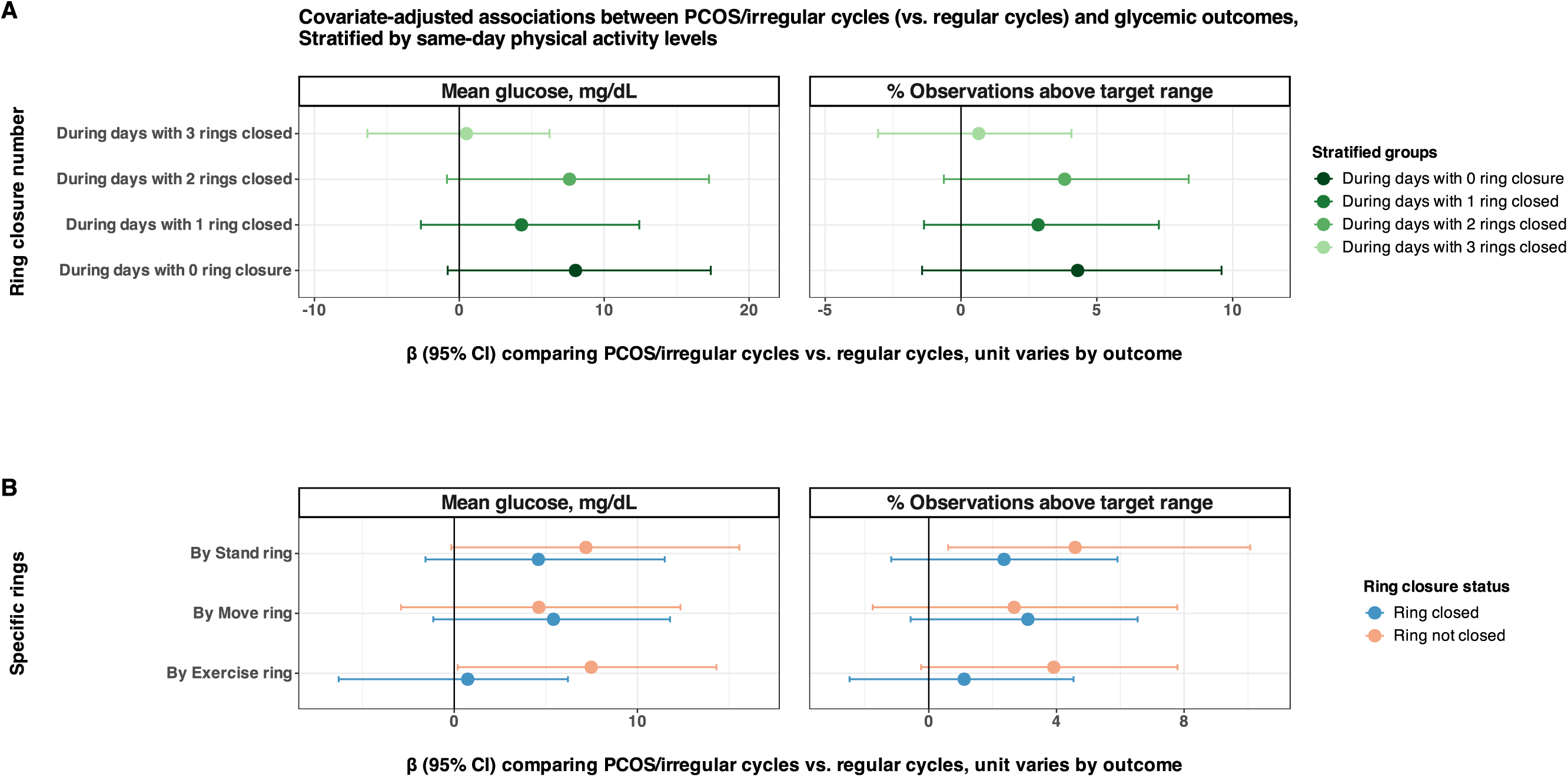
Associations between PCOS/EL irregularity and mean glucose stratified by same-day physical activity, among a subset of 521 participants with eligible 113,324 days of Apple Watch data during CGM monitoring. (A) Covariate-adjusted associations stratified by same-day number of rings closed; (B) Covariate-adjusted associations stratified by specific ring (move, exercise, stand) closure. The three rings in Apple Watch include: Move (active calories burned), Exercise (minutes), and Stand (hours); Daily ring closure for each ring is defined as meeting or exceeding self-set personal goals. Number of person-days in each number of ring closure is summarized in Table 1. Models adjusted for age, self-reported prediabetes, type 1 diabetes, type 2 diabetes, BMI, education level, race/ethnicity, socioeconomic status, typical physical activity level, typical low carb diet preference, and stress reflected by PSS-4 score. All covariates are self-reported at enrollment. Bias-corrected bootstrap confidence intervals were estimated from 1000 bootstrap samples.

First, among 521 participants (who provided consent and opted in to share relevant physical activity data with the Study) with 113,324 days of wearable-derived same-day physical activity data during glucose monitoring (Figure 3, estimates and 95% CIs detailed Supplemental Table S3), the magnitude of associations between having PCOS/PMOS or irregular cycles and higher glucose or higher %observations above target range both showed suggestive attenuation during days when all three physical activity rings (move, exercise, stand) were closed (i.e., when meeting all self-set targets). When evaluating specific ring closure status (Figure 3B), the magnitude of associations between having PCOS/PMOS or irregular cycles and higher glucose levels showed an attenuation to almost zero during days when the exercise ring was closed (i.e., when meeting daily targets for exercise minutes); smaller-magnitude attenuation was observed for closing the stand ring (i.e., when meeting daily targets for stand hours), but not for the move ring (i.e., when meeting daily targets for total calories burned).

Second, among 58 participants with 2,565 days of same-day dietary logs (who provided consent and opted in to share relevant dietary log data with the Study) during glucose monitoring (Figure 4), the magnitude of associations between having PCOS/PMOS or irregular cycles and higher glucose levels appeared larger during days characterized by diets rich in refined carbohydrates, with higher relative carbohydrate intake, or with lower relative protein intake, whereas days with dietary compositions generally known to be beneficial for glucose management (e.g., diet rich in fiber, with lower relative carbohydrate intake, or with higher relative protein intake) had substantially smaller (close to or below zero) glycemic differences between the PCOS/PMOS or irregular cycles group vs. the regular cycles group.

**Figure 4.**
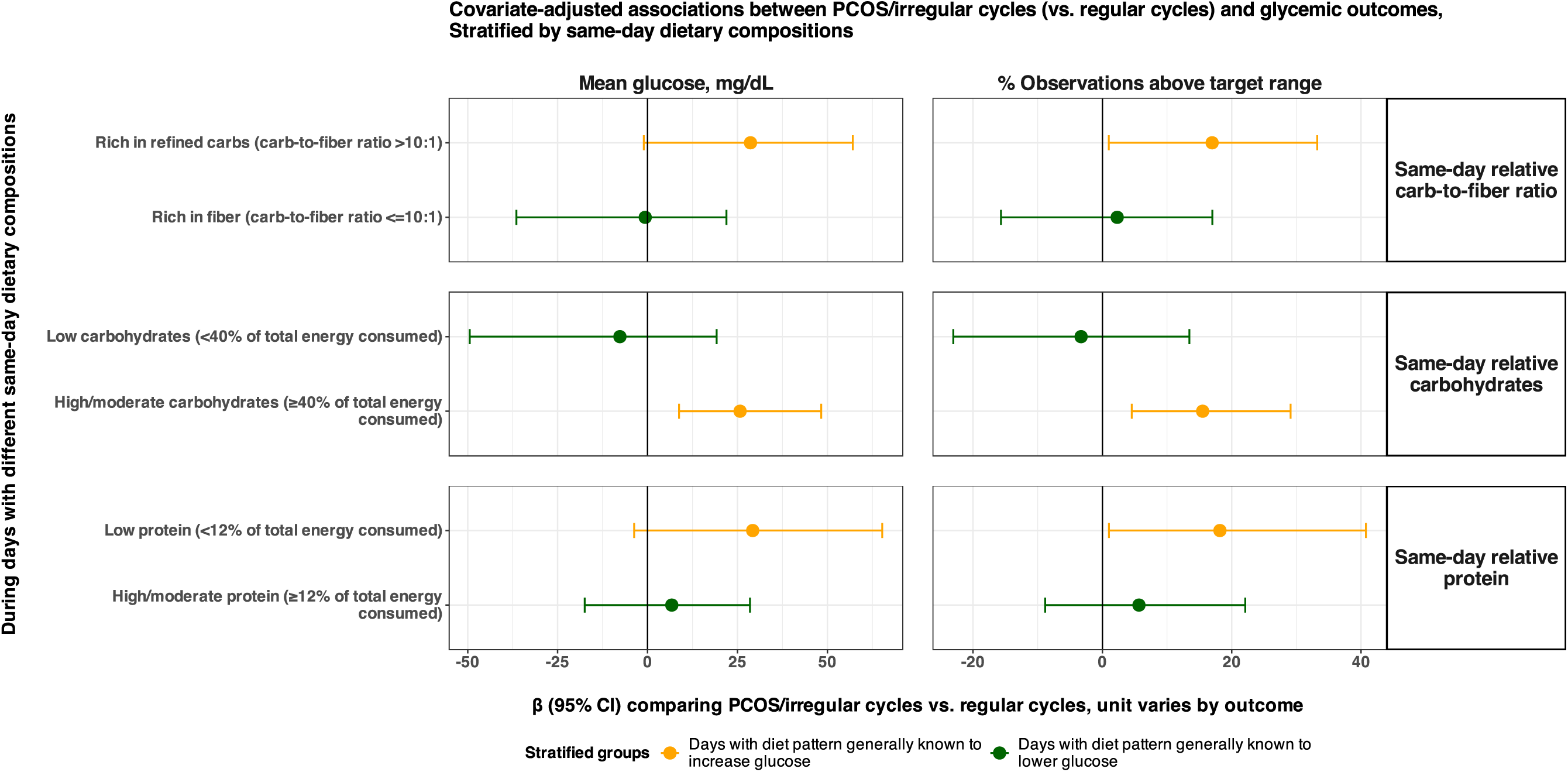
Associations between PCOS/EL irregularity and mean glucose stratified by same-day dietary composition, among a subset of 58 participants with digitally logged dietary data across 2,565 days during CGM monitoring. Models adjusted for age, self-reported prediabetes, type 1 diabetes, type 2 diabetes, BMI, education level, race/ethnicity, socioeconomic status, typical physical activity level, typical low carb diet preference, and stress reflected by PSS-4 score. All covariates are self-reported at enrollment. Bias-corrected bootstrap confidence intervals were estimated from 1000 bootstrap samples.

Lastly, suggestive modifications by common diabetes medications are observed. Among 180 participants who provided consent and opted in to share their EHR medication records across 65,885 days during glucose monitoring (Figure 5), the association between PCOS/PMOS or irregular cycles and mean daily glucose was 17.27 mg/dL (95% CI: 0.13, 33.32) among person-days when no common diabetes medications were used. In contrast, the point estimates for the associations attenuated to 3.29 mg/dL among person-days when any common diabetes medication was used, and to 9.68 and 8.29 mg/dL among specific person-days when metformin or GLP-1 RAs were used, respectively.

**Figure 5.**
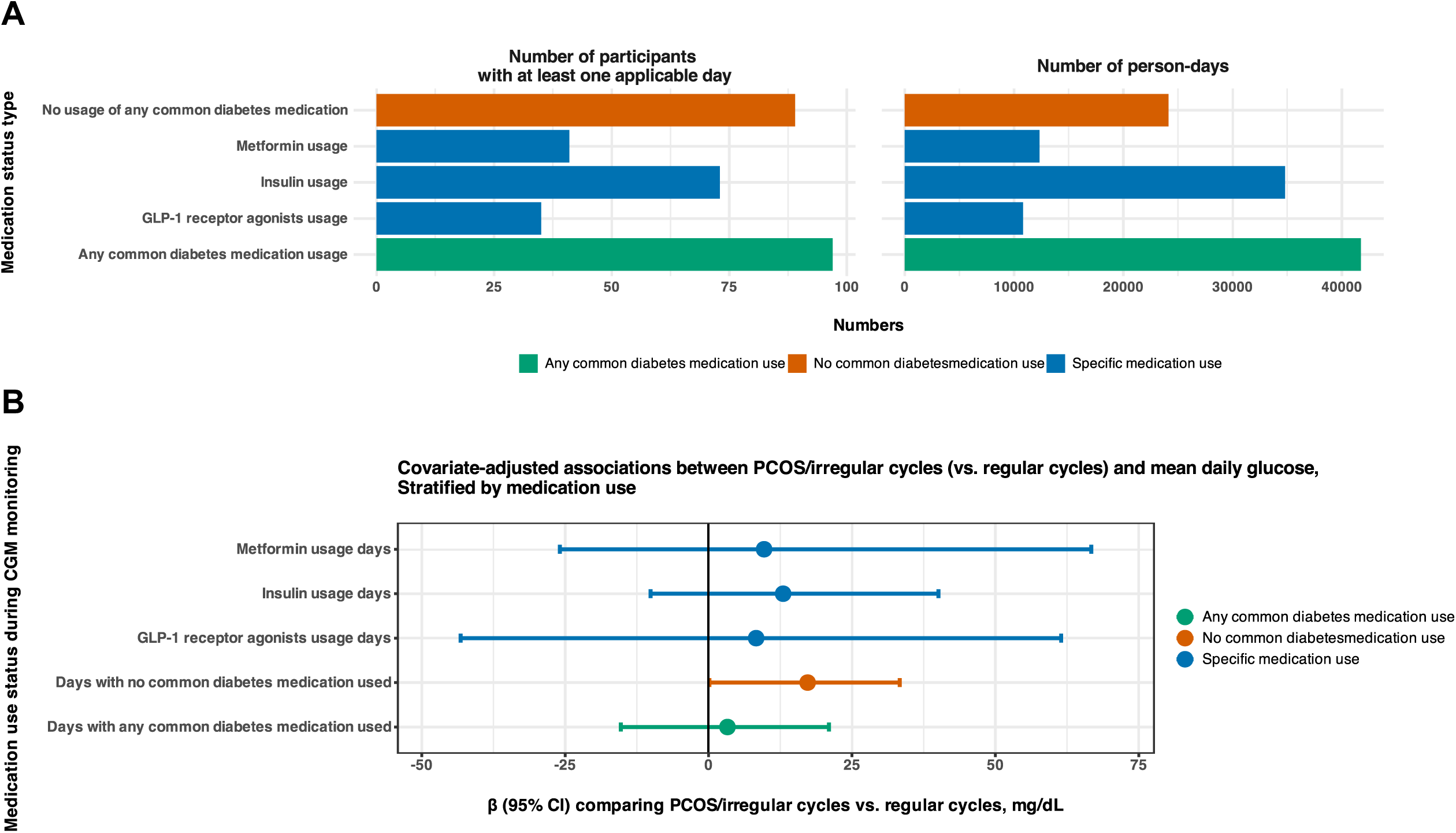
Associations between PCOS/EL irregularity and mean glucose stratified by medication use from shared health records, among a subset of 180 participants with 65,885 days during CGM monitoring. (A) Number of participants and person-days; (B) Covariate-adjusted association estimates (95% CIs) stratified by medication use. Number of participants applicable in each category may sum more than the total number of participants, as a person could have days with medication use while other days without medication use, and they could have taken ≥2 medications on a given day. Models adjusted for age, self-reported prediabetes, type 1 diabetes, type 2 diabetes, BMI, education level, race/ethnicity, socioeconomic status, typical physical activity level, typical low carb diet preference, and stress reflected by PSS-4 score. All covariates are self-reported at enrollment. Bias-corrected bootstrap confidence intervals were estimated from 1000 bootstrap samples.

### Secondary analysis results: Associations stratified by preexisting diabetes status

Supplemental Table S4 shows the covariate-adjusted exposure-outcome associations, separately among participants without any self-reported diagnosis of type 1 or type 2 diabetes, participants with type 1 diabetes, and participants with type 2 diabetes (see distributions in Table 1). Positive associations between PCOS/PMOS or early-life irregular cycles and higher glucose or higher %observations above target range remained among participants without any self-reported diagnosis of type 1 or type 2 diabetes, and among participants with type 1 diabetes. Among participants with type 2 diabetes, higher glycemic measures were only suggested in the early-life irregular cycles, but not in the PCOS/PMOS group.

### Secondary analysis results: PCOS/PMOS by androgen excess symptoms

A subset of 661 participants responded to the relevant Hormonal Symptoms Survey that had information about the presence or absence of androgen excess symptoms. Imputed to all 716 participants using multiple imputations, for individuals with PCOS/PMOS compared to the referent group, the magnitude of associations with higher mean glucose and higher %observations above target range were larger among participants who reported signs of potential androgen excess (self-reported hirsutism and/or moderate/severe acne)^18^; in contrast the group with PCOS/PMOS and without self-reported androgen excess symptoms showed suggestive associations with higher glucose variability reflected by the standard deviation values (Supplemental Table S5).

### Sensitivity analysis results

As an exploration of potential physiological cardiometabolic response to elevated glucose, we observed a potential correlation between higher daily mean glucose and higher daily mean resting heart rate (RHR) among participants who provided consent and opted in to share relevant RHR data with the Study, as shown in Supplemental Figure S3. Across the range of mean daily glucose values, mean RHR had an increasing trend as glucose levels increase, with similar trends observed among the three exposure groups, while the PCOS/PMOS group had the highest absolute RHR levels compared to the other two groups.

Additionally, unadjusted aggregated hourly data across a 24-hour window show that individuals with irregular cycles or PCOS/PMOS had consistently higher mean glucose compared to the regular cycles group across all hours of the day (Supplemental Figure S4). Despite differences in absolute values, all three groups had similar relative changes regarding expected circadian glucose variation, with all exposure groups having peak values during mid-afternoon (around 2-3pm) and evening hours (around 8-9pm), and lowest values during early morning hours (around 6-7am).

For the three categories of potential modifiers (physical activity, diet, or medication use), correlations are visualized among subsets of participants where more than one category’s data is available (Supplemental Figures S5, S6, S7). Activity ring closures were highly-to moderately correlated amongst different rings, and moderate negative correlations were seen for relative carbohydrate intake vs. relative protein intake; however, correlations between physical activity and diet were relatively weak, with the highest correlation being between number of activity ring closed and carbohydrate-to-fiber intake ratio (a negative correlation of −0.26). Use of metformin and GLP-1 RAs were moderately correlated, but their correlations with activity ring or dietary patterns were very weak.

In sensitivity analyses further adjusted for ever hormone use, the effect estimates for PCOS/PMOS or irregular cycles and mean glucose were similar to that in the main analyses [β (95% CI): 6.77 (−0.95, 14.47) mg/dL for PCOS/PMOS, 6.31 (−1.33, 14.49) mg/dL for irregular cycles]. In sensitivity analyses removing 17 participants who reported being pregnant during any time period of their glucose monitoring, the effect estimates for PCOS/PMOS or irregular cycles and mean glucose were similar to that in the main analyses [β (95% CI): 6.86 (−0.66, 14.68) mg/dL for PCOS/PMOS, 6.27 (−1.26, 14.40) mg/dL for irregular cycles].

## Discussion

Within a US digital cohort, by integrating longitudinally assessed CGM data, digital survey responses, wearable-derived physical activity metrics, dietary logs, and linked clinical information, we observed less favorable glycemic patterns across multiple CGM-derived outcomes associated with having PCOS/PMOS or early-life irregular cycles, including higher mean glucose level, lower percentage of observations in target range and higher percentage of observations above target range. Participants with PCOS/PMOS who have androgen excess symptoms were most impacted by the higher glucose patterns. Higher physical activity (i.e., meeting self-set activity goals), glucose-lowering diet patterns (characterized by relatively lower carbohydrates, higher fiber, or higher protein), or use of certain medications (e.g., metformin or GLP-1 RAs) have the potential to offset the glycemic risks associated with PCOS/PMOS or menstrual irregularity. While further validations in other populations are needed, these findings suggest that reproductive health phenotypes within and beyond clinically diagnosed PCOS/PMOS may capture broader dimensions of glycemic and metabolic vulnerability, and may have the potential to be used as an early vital sign to identify women at elevated glycemic risk. Lifestyle modification findings also provide potential personalized intervention opportunities.

Our findings align with existing literature that found an association between PCOS/PMOS and insulin resistance, impaired glucose metabolism, and elevated long-term risk of type 2 diabetes and cardiovascular disease^3–5^. Much of the prior literature has relied on traditional clinical assessments or biomarkers (e.g., fasting glucose, HbA1c, or oral glucose tolerance test). In contrast, CGM-derived metrics provide a more temporally refined characterization of glucose dynamics in real-world settings, enabling assessment not only for within-person average glucose levels, but also daily variability and time spent within clinically relevant glycemic ranges. By analyzing CGM-derived outcomes, our study of 716 individuals with varying menstrual health and general health phenotypes extends beyond small pilot studies that evaluated CGM outcomes in relation to clinically diagnosed PCOS/PMOS^12–15^. In particular, we observed associations of early-life menstrual irregularity, in the absence of diagnosed PCOS/PMOS, with higher glucose levels, lower %observations in target range, higher %observations above target range, and higher GMI derived from CGM values. Menstrual irregularity has historically been conceptualized primarily within reproductive contexts, but growing evidence now suggests that menstrual cycle characteristics may also reflect broader endocrine and metabolic risks over the life course^19,20^. Our findings from CGM data support the hypothesis that menstrual irregularity may serve as an early-life marker of future metabolic risks, even among individuals who do not meet diagnostic criteria for PCOS/PMOS or do not have access to receive a PCOS/PMOS diagnosis. Additionally, the associations were generally weaker for the irregular cycle group than the PCOS/PMOS group. This suggests the possibility of a continuum of metabolic risk with the irregular cycle group exhibiting possible early life metabolic dysregulation, while the PCOS/PMOS group may have more established insulin resistance and chronic high glucose, with lower variability. Rizzi et al. observed lower glucose variability among patients with PCOS/PMOS and insulin resistance compared to patients with PCOS/PMOS and without insulin resistance^14^, suggesting future studies with data on whether someone already established insulin resistance is needed to understand glucose variability. Combined with GMI and lab HbA1c data, the increase of around 0.4 %hemoglobin suggests that our findings are clinically relevant, as compared to the 0.3 %hemoglobin threshold for evaluating effectiveness of glucose lowering therapies^21^. Another study that examined the clinically relevant increase in macro- and micro-vascular complications being associated with 1 %hemoglobin higher HbA1c^22^ is consistent with the exploratory signals we observed for elevated resting heart rate. These findings suggest that both early-life menstrual irregularity and PCOS/PMOS may influence real-world glycemic patterns that may carry lifelong cardiometabolic health implications.

Using same-day wearable-derived physical activity data, we found that daily physical activity appeared to attenuate glycemic differences associated with PCOS and menstrual irregularity, with the largest glycemic differences observed during low activity days and the smallest during days when all personal activity goals are met. The trend towards reduction also appeared to be most prominent among meeting personal exercise minutes goals, compared to stand or total calories goals. While these observational findings in small subsets may not establish causality, this preliminary evidence suggests that day-to-day physical activity behaviors, especially intentional exercise goals, may provide a partial buffer for the metabolic risks among individuals with PCOS/PMOS or irregular cycles compared to their peers with regular cycles. Same-day dietary composition also showed potential modification for the associations between PCOS/PMOS or irregular cycles with glucose, with larger glucose differences during days with diets rich in refined carbohydrates, higher relative carbohydrate intake, or lower relative protein intake, whereas dietary patterns commonly perceived as beneficial for glucose management were associated with attenuated glycemic differences. By integrating CGM data with same-day dietary logs, these findings support a framework for developing more personalized dietary recommendations based on individual physiologic responses and reproductive health phenotypes^23^. Lastly, potential effect modification by medications such as metformin and GLP-1 RAs is consistent with their primary indication for treating diabetes. Given recent uptake of GLP-1 RAs use among individuals without diabetes, including individuals impacted by PCOS/PMOS or menstrual irregularity^24,25^, future studies with larger sample size of medication use data may help clarify the extent to which medications alter glycemic dynamics in these populations.

While there was higher mean glucose but smaller glucose variability observed in the PCOS/PMOS group with androgen excess, there was lower mean glucose but higher variability in the PCOS/PMOS group without andron excess, which may suggest the latter phenotype being prone to less sustained insulin resistance but more volatile glucose patterns. In contrast, PCOS/PMOS with androgen excess may be characterized by more persistent metabolic insulin resistance relevant to the role of androgen^26^, resulting in higher but relatively stable glucose profiles. However, these potential mechanisms are not confirmed in this study, as measures of insulin sensitivity or meal-specific glucose excursions were not directly measured.

In stratified analyses, we found that the association between PCOS/PMOS and higher glucose levels was most pronounced among participants without diabetes, or those with type 1 diabetes, but was attenuated in participants with type 2 diabetes. Our hypothesis was that individuals with both PCOS/PMOS and type 2 diabetes may be a sub-population already prescribed with intensive lifestyle and medical interventions targeting glucose control, potentially masking the PCOS/PMOS-specific metabolic effects. The remained positive associations between PCOS/PMOS and higher glucose levels in those without a diabetes diagnosis suggest the potential role of PCOS/PMOS in increasing glycemic risk even among relatively metabolically healthy women, highlighting the need for early risk identification among broader populations. Type 1 diabetes is overrepresented in our data, which reflects real-world scenarios where a CGM device is considered standard of care for type 1 diabetes management that requires insulin. This differential uptake of CGM technology between diabetes types may have selected for participants with type 1 diabetes who may have greater engagement in metabolic self-monitoring in our data.

One limitation of this study is that PCOS/PMOS diagnosis, menstrual cycle irregularity, and daily dietary logs were based on self-reports and may be subject to misclassification. Early-life menstrual irregularity may reflect heterogeneous underlying etiologies (e.g., other endocrinopathies) that could not be further distinguished in this cohort. Behavioral measures like physical activity ring closure, although passively collected from wearables with high temporal resolution, may not fully capture true activity levels on the absolute scale. Despite adjustment for multiple covariates, residual or unmeasured confounding is still possible, especially if there are any time-varying behavioral factors associated with daily variations in glucose patterns. All participants with diabetes diagnosis had their earliest CGM reading after receiving the diagnosis; and 99.6% of the participants with PCOS/PMOS had their earliest CGM reading after receiving PCOS/PMOS diagnosis. As such, while reverse causality cannot be fully ruled out, we believe the risk is low. We also acknowledge potential confounding by indication for the associations observed among medication users, where participants with more severe metabolic dysfunction or PCOS/PMOS-related hyperandrogenism may have preferentially received metformin or GLP-1 RAs, which may lead to observed findings reflecting a mix of underlying metabolic severity and true medication benefits. Lastly, given the study’s inclusion criteria, our findings may not be fully generalizable to all US individuals who menstruate or to other populations. Selection bias may have occurred in the effect modification analyses which used subsets with relevant wearables, digital log, or clinical data.

Our study has several unique strengths. First, we integrated multiple, complementary digital health data sources, including CGM measurements, wearable-derived physical activity, digital dietary logs, and linked health records, enabling a comprehensive evaluation of not only CGM metrics associated with different health phenotypes, but also the roles of real-world lifestyle factors and medications. Second, use of CGM-derived outcomes from dense, longitudinal data enabled us to both validate against and characterize beyond sparse, traditional laboratory biomarkers. Third, evaluation of same-day behavioral effect modification provides real-time insight into how lifestyle factors may interact with underlying menstrual health phenotypes on a daily basis. Fourth, the evaluation of PCOS/PMOS, menstrual irregularity independent of PCOS/PMOS, and the sensitivity analyses further stratifying by androgen excess symptoms broadens the understanding of metabolic risks beyond traditional diagnostic categories.

Overall, our study suggested that PCOS/PMOS and early-life irregular cycles may be associated with elevated glucose and other glycemic related risks. Meeting daily personal physical activity goals, consuming diet that’s rich in fiber or protein, and using specific medications (e.g., GLP-1 or metformin) may have the potential to offset these risks. The integration of CGM device data, wearables, digital dietary logging, and linked health records provides a unique opportunity to characterize glycemic characteristics at daily levels, and to potentially identify individuals at elevated metabolic risk early during the life course, offering potentials for personalized metabolic screening and modifications.

## Methods

### Study design and population

This analysis used data from the Apple Women’s Health Study (AWHS), a prospective digital cohort study in the United States designed to characterize menstrual cycles, health and behavior factors. AWHS eligibility criteria include being user of the Apple Research app on their iPhone (sole usage, including their iCloud account), have ever menstruated, live in the United States, being at least 18 years old (19 in Alabama and Nebraska, 21 in Puerto Rico), and able to communicate in English. The AWHS was approved by the institutional review board at Advarra (CIRB #PRO00037562). Enrollment began in November 2019 and is ongoing. Written informed consent of participation was provided at enrollment. More information of the AWHS study design, eligibility and data were described previously^27,28^.

For this analysis, we included a subset of AWHS participants who enrolled from 11/2019 to 08/2026, provided responses to questions on demographics, medical history, and reproductive health surveys, and shared with this study any glucose data logged or synced from any third-party apps or devices in the Apple Health app. To enhance data quality, we restricted our analysis to eligible days with confirmed CGM device information (99.6% from Dexcom G6 and 0.4% from FreeStyle Libre 3) and a minimum of 140 daily readings (i.e., at least half of the expected ∼280 daily readings from approximately 5-minute measurement intervals). We further restricted to participants with at least seven eligible CGM days. Dexcom G6 and FreeStyle Libre 3 do not require fingerstick calibrations, although voluntary calibration is possible; the seven-day minimum was intended to minimize any potential, voluntary calibration effects and enhance data reliability to reflect usual patterns^29^. A flowchart is available in Supplemental Figure S1. A summary of data sources of key variables is shown in Supplemental Table S6. Participants in the Study who provided data for this analysis provided their consent and opted in to share relevant data with the Study.

### PCOS/PMOS and early-life menstrual irregularity

We defined three mutually exclusive exposure groups based on survey responses at enrollment. The first group (**PCOS/PMOS**) included participants who self-reported having been diagnosed with PCOS “by a doctor or other care provider”. Among participants who did not report a PCOS/PMOS diagnosis, the second group (**irregular cycles**) included those who reported not spontaneously developing regular cycles within 4 years after menarche or used hormones to achieve regularity; the third group (**regular cycles**, as the referent group) were those who spontaneously established regular cycles within 4 years of menarche. Participants with missing responses (<0.5%) were excluded from the analytical dataset (Supplemental Figure S1).

### Glycemic outcomes

For each day per each participant with eligible CGM readings, we calculated the following primary outcomes for daily glucose: mean, standard deviation (SD), coefficient of variation (CV), and percentages of CGM observations above target range (>180 mg/dL), below target range (<70 mg/dL), and in target range (70–180 mg/dL)^30^ based on observed CGM data on valid days. These metrics reflect overall glucose levels, variability, and glycemic control.

As secondary analyses, we evaluated the glucose management indicator (GMI), calculated as GMI (%) = 3.31 + 0.02392*[mean glucose in mg/dL], serving as an approximation of HbA1c levels^31^. We also evaluated direct HbA1c lab results from a subset of study participants who shared their electronic health records.

### Covariates

We considered the following self-reported variables at enrollment as potential confounders of the exposure group-glycemic outcome associations: age (18–29, 30–39, 40–49, or ≥50 years), physician-diagnosed prediabetes, type 1 diabetes, or type 2 diabetes, body mass index (BMI) (<25, 25-29.9, or ≥30 kg/m^2^), race/ethnicity [non-Hispanic White, non-Hispanic Black, Hispanic, or multiple/other (Asian, American Indian or Alaska Native, Middle Eastern or North African, Native Hawaiian or Pacific Islander, or “None of these fully describe me”)], education (≤high school, some college or tech school, college degree, or graduate school), subjective MacArthur Scale of Subjective Social Status ^32^ (high vs. medium/low), and physical activity levels (typical exercise time >75 vs. ≤75 minutes per week), typical pattern of low carbohydrates diet (yes/no), and stress reflected by the Perceived Stress Scale 4 (PSS-4) (standard values from 0 to 16).

### Effect modifiers

We evaluated the following factors as potential effect modifiers: physical activity patterns from wearables, dietary macronutrient composition from digital food logs, and diabetes-relevant medication use from linkage of health records. These factors were selected as they are known to influence glucose control^33,34^. We hypothesized that these potentially modifiable lifestyle/medication factors may attenuate partly the adverse glycemic outcomes associated with PCOS/PMOS or menstrual irregularity, by interacting with the underlying pathophysiologic mechanisms^35,36^. Secondary analyses of these factors as effect modifiers, detailed below, were restricted to subsets of participants with available data for each variable, with sample sizes for each of these secondary analyses provided below. As these subsets are small-to-moderate in size, we dichotomized the exposure groups into PCOS/PMOS and/or irregular cycles (a group prone to irregular cycles) vs. regular cycles (the referent group).

#### Effect modifier: same-day physical activity patterns

Daily physical activity (PA) was inferred from Apple Watch activity ring metrics [Move (active energy burned), Exercise (intentionally logged exercise minutes), and Stand (hours with ≥1 full minute of movement)] among a subset of 521 participants who shared this data with the study, restricted to days with at least 14 hours of Apple Watch wear. Descriptions of these sensor-based metrics can be found in previous publications based on the same or similar cohorts^37,38^. We used daily number of ring closures (0, 1, 2, or 3) or whether closing each specific ring for each person-day with both ring closure and CGM data (113,324 days in total), to evaluate goal-oriented PA as a potential modifier for the association between PCOS/PMOS or early-life irregularity and glucose levels.

#### Effect modifier: dietary macronutrient patterns

Daily dietary composition was assessed among 58 participants who shared dietary macronutrient data (Apple Health app or other third-party apps that the participant allows to write to the Health app), contributing 2,565 days with same-day CGM data and select macronutrient data, including daily sum of total energy consumed (kCal), and carbohydrates, fiber, sugar, fat, and protein (grams). We derived three relevant daily relative macronutrient composition measures as potential effect modifiers for the PCOS/PMOS or early-life irregularity and glucose levels association: 1) relative carbohydrates: high/moderate (≥40%) vs. low (<40%) of total energy consumed; 2) carbs-to-fiber ratio: >10:1 (rich in refined carbohydrates) vs. ≤10:1 (rich in fiber); and 3) relative protein: high (≥12%) vs. low (<12%) of total energy consumed.

#### Effect modifier: medication use

Medication use was extracted from electronic health records (EHR) among 180 participants who linked their EHR data, shared at least one medication use record, and had available CGM data for this analysis. We screened for common medication types for diabetes [metformin, glucagon-like peptide-1 receptor agonists (GLP-1 RAs), insulin, dipeptidyl peptidase-4 (DPP-4) inhibitors, sodium-glucose cotransporter-2 (SGLT2) inhibitors, sulfonylureas, thiazolidinediones, or other medications like alpha-glucosidase inhibitors, or meglitinides] and evaluated the irregular cycles and glucose levels associations in the following groups: not using any common diabetes medications during person-days with CGM data, or using any common diabetes medications during person-days with CGM data [and further investigated subgroups specifically using metformin, glucagon-like peptide-1 receptor agonists (GLP-1 RAs), or insulin].

### Statistical analysis

We summarized participant characteristics, CGM data density, and glycemic measures overall and by exposure groups using descriptive statistics. To evaluate the associations between exposure groups and each daily average glycemic measures, we used cluster-weighted generalized estimating equations (GEE), with weights defined as the inverse of the cluster size (i.e. the inverse of the number of days a given participant contributes to the analysis)^39^, adjusting for abovementioned covariates. We computed 95% confidence intervals (CIs) using nonparametric bootstrap with 1,000 resamples, resampling at the participant level with replacement. Bias-corrected and accelerated (BCa) 95% CIs were derived^40^.

As secondary analyses, we explored potential effect modification by physical activity, dietary composition, and medication use as detailed above. Given small-to-moderate sizes of these subsets, we performed exploratory stratified analyses and examined the magnitude and direction of associations across strata, as opposed to evaluating formal tests of interaction.

In other sensitivity analyses, we: (1) further classified the PCOS/PMOS group into PCOS/PMOS with androgen excess (AE) symptoms (self-reported signs of hirsutism or moderate/severe acne^41^) and PCOS/PMOS without AE symptoms; (2) evaluated differences in Apple Watch measured resting heart rate (RHR) as a relevant cardiometabolic vital sign in relation to elevated glucose; (3) summarized aggregated 24-hour glucose levels by each hour to further explore hourly patterns; (4) evaluated the robustness of main findings after further adjusting for ever hormone use (mainly exogenous hormonal contraceptives or hormones for menopause); and (5) evaluated the associations after excluding 17 participants who had any CGM measurements that occurred during a pregnancy window to remove any potential impact from pregnancy (as pregnancy is considered as sensitive window with altered glucose metabolism adaptations)^42^.

All analyses were conducted in Python (version 3.6) and R (version 4.1.2). Effect estimates were accompanied by 95% confidence intervals (CIs).

## Data availability

Aggregated data that support the findings of this study may be available upon reasonable request from the corresponding author and senior author. Any request for data will be evaluated and responded to in a manner consistent with legal obligation and policies intended to protect participant confidentiality, language in the Study protocol, and informed consent form.

## Acknowledgements

We would like to thank the study participants for consenting and contributing to the advancement of women’s health research. We would also like to acknowledge previous AWHS staff at Harvard T.H. Chan School of Public Health (Carrie Sarcione, Erin Dracup, Eliana Huffman) for their work in supporting this study, and Dr. Anne Marie Jukic and Dr. Donna D. Baird for their contributions to the preliminary abstract presented at Endocrine Society’s ENDO 2026 in June 2026.

## Author contributions

Z.W. was responsible for study design, data preparation, statistical analyses, interpretation of the results, manuscript writing, revision, and finalization. G.A. contributed to study design, methodology, interpretation of the results, and review of the preliminary work. B.Z. contributed to part of the data preparation, interpretation of the results, and manuscript revision. S.W., P.B-R., J-P.O., M.A.W., and B.A.C. provided critical reviews to the analysis, contributed to the interpretation of the results, and contributed to manuscript revision. S.S. contributed to review of content in relation to the publication policy of Apple Inc, and contributed to manuscript revision. S.M. oversaw all aspects of the work and contributed to providing critical reviews to the study design and analysis, interpretation of the results, and manuscript revision.

## Prior conference proceedings

A preliminary version of this work was presented on June 14, 2026 at the Endocrine Society’s ENDO2026 during June 13-16, 2026, in Chicago, IL, as a poster presentation. Correspondingly, the accepted preliminary abstract at ENDO2026 is expected to be published in a supplemental issue in the *Journal of the Endocrine Society (JES) as “ENDO 2026 Abstracts Annual Meeting of the Endocrine Society”.* This manuscript submission has a substantial extension of results, methodology, analysis, conclusions and/or implications compared to the conference abstract.

## Study registration

This study is not a clinical trial. However, the parent cohort, Apple Women’s Health Study, is registered on Clinicaltrials.gov (NCT04196595): https://classic.clinicaltrials.gov/ct2/show/NCT04196595

