## Supplemental for "Glycemic patterns from continuous glucose monitoring devices among women with regular and irregular menstrual cycles"

**Supplemental Materials**

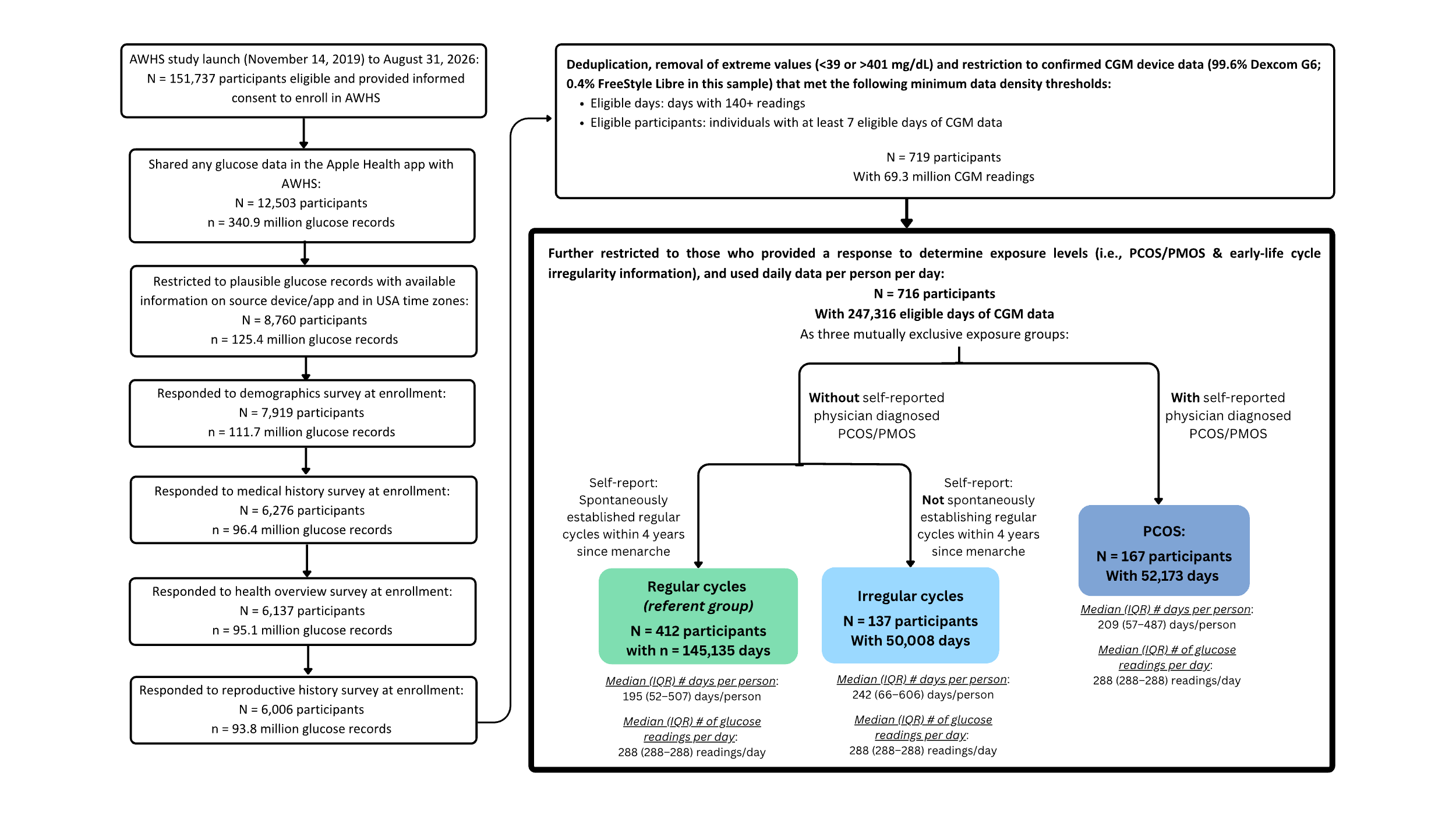

### **Supplemental Figure S1.** Participant flowchart for the main analyses.

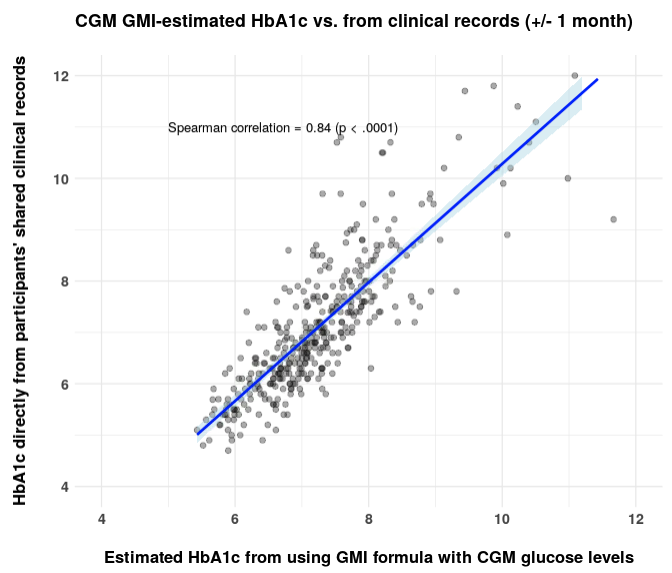

**Supplemental Figure S2.** Correlations between GMI-estimated HbA1c from CGM data and HbA1c values directly from clinical records, among the subset of 139 participants with clinical HbA1c lab records within +/- 1 month around any of their CGM readings.

GMI estimated HbA1c (%) = 3.31 + 0.02392 × [mean glucose in mg/dL] (Bergenstal et al. 2018. *Diabetes Care*)

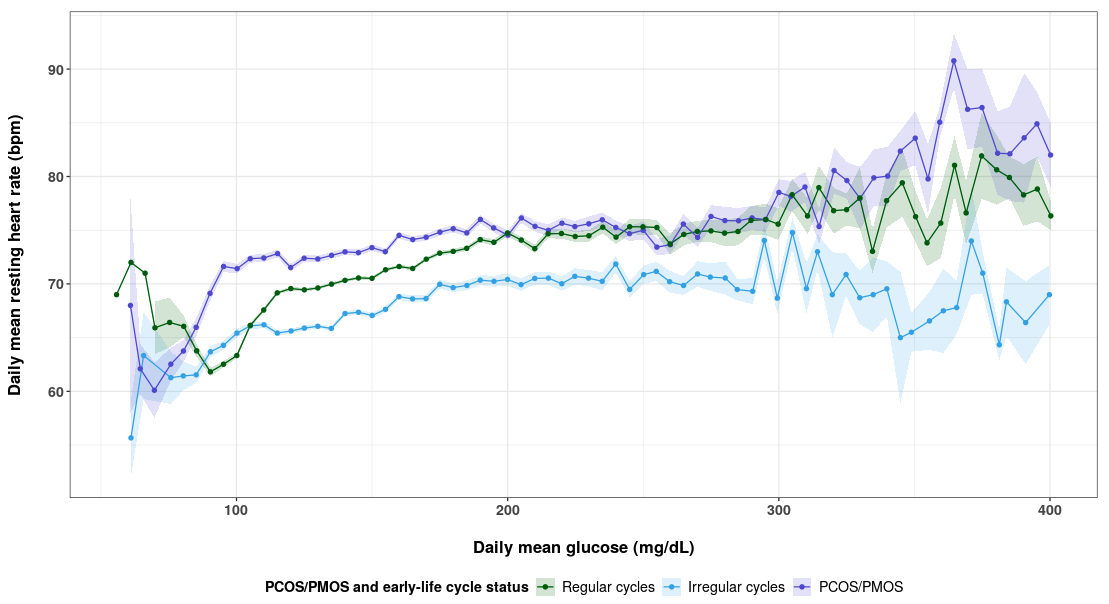

**Supplemental Figure S3.** Daily mean glucose from CGM and daily mean resting heart rate from Apple Watch.

Aggregated data shown, not adjusted for any covariates.

Abbreviations: CGM, continuous glucose monitoring; PCOS, polycystic ovary syndrome [in May 2026, PCOS was renamed as polyendocrine metabolic ovarian syndrome (PMOS)]. Regular cycles group: those without self-reported PCOS diagnosis, and reported spontaneously establishing regular cycles within 4 years after menarche; Irregular cycles group: those without self-reported PCOS diagnosis, and reported reaching cycle regularity in 5+ years after menarche, their cycles are still irregular, or used hormones to achieve regularity.

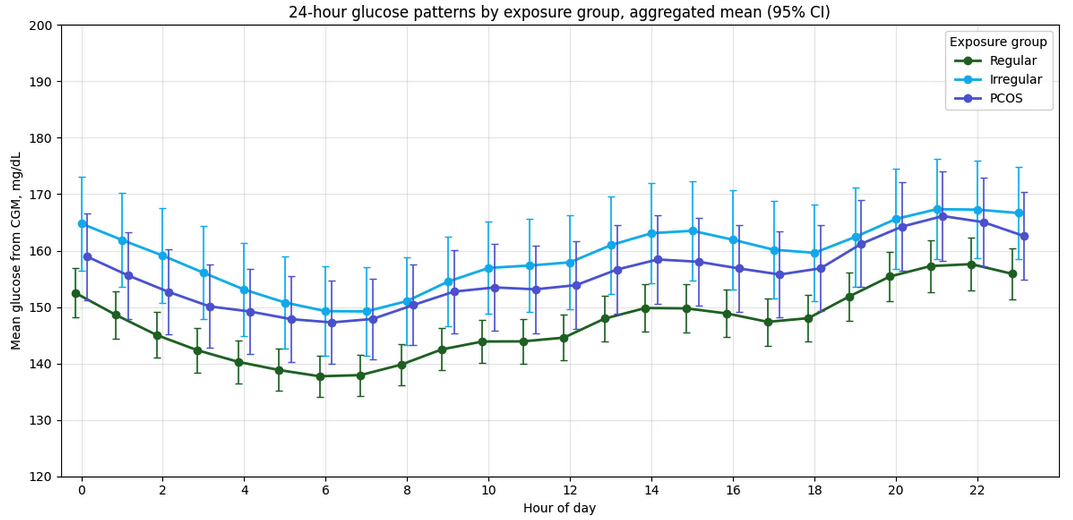

**Supplemental Figure S4.** Aggregated 24-hour glucose patterns by exposure groups.

Aggregated data shown across all relevant CGM readings in each group, not adjusted for any covariates.

PCOS, polycystic ovary syndrome. In May 2026, PCOS was renamed as polyendocrine metabolic ovarian syndrome (PMOS). We use PCOS throughout this manuscript because it was the term used when survey data were collected, prior to the May 2026 renaming.

Regular cycles group: those without self-reported PCOS/PMOS diagnosis, and reported spontaneously establishing regular cycles within 4 years after menarche; Irregular cycles group: those without self-reported PCOS/PMOS diagnosis, and reported reaching cycle regularity in 5+ years after menarche, their cycles are still irregular, or used hormones to achieve regularity.

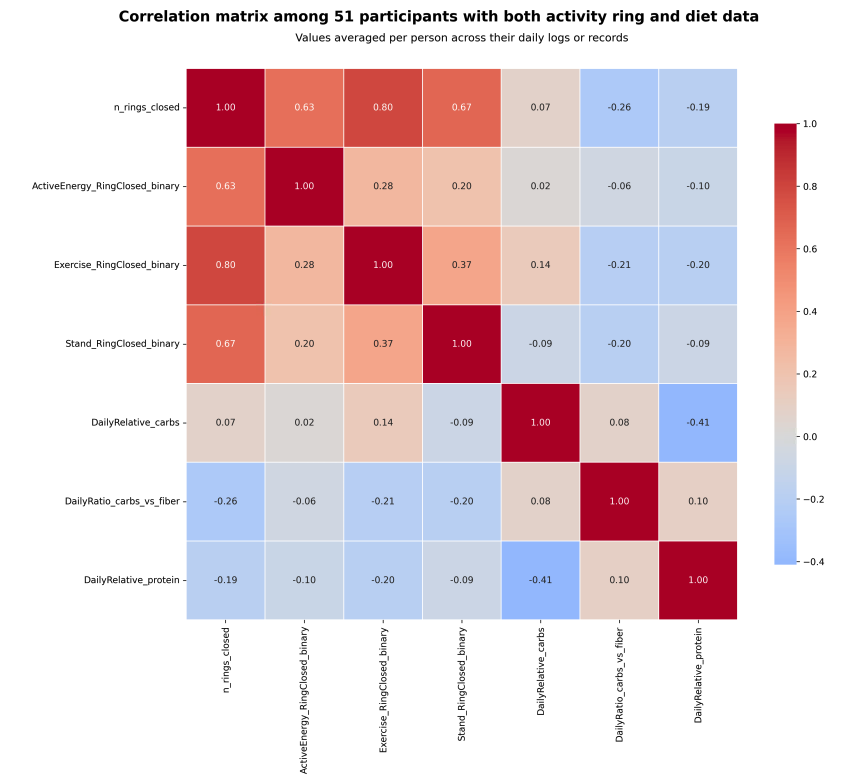

**Supplemental Figure S5.** Spearman correlations for effect modifiers, among 51 participants with both activity ring and self-logged diet data.

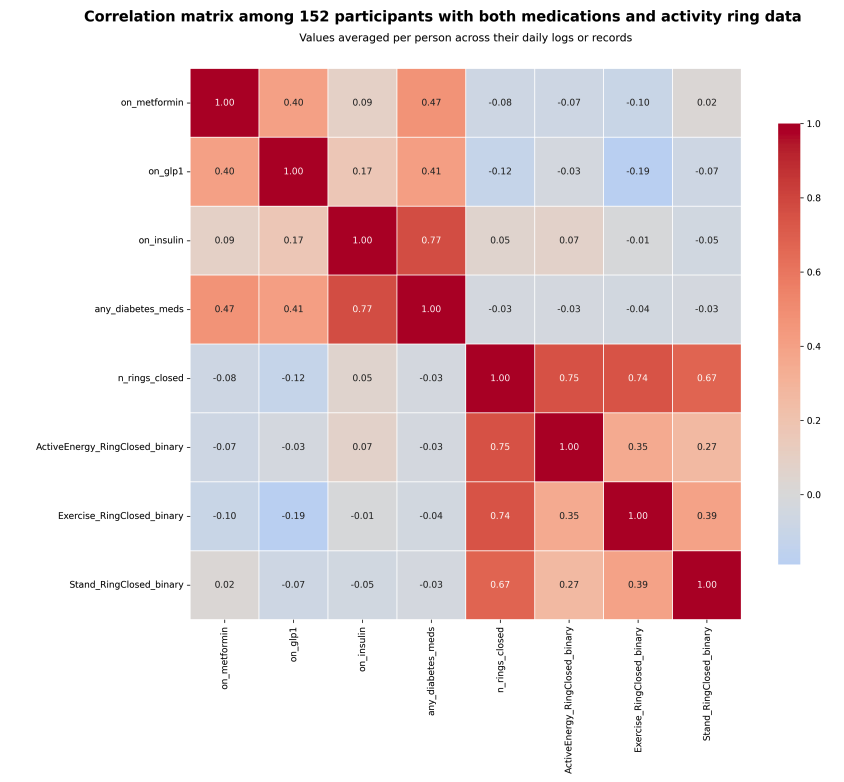

**Supplemental Figure S6.** Spearman correlations for effect modifiers, among 152 participants with both wearable-based activity ring and EHR-based medications data.

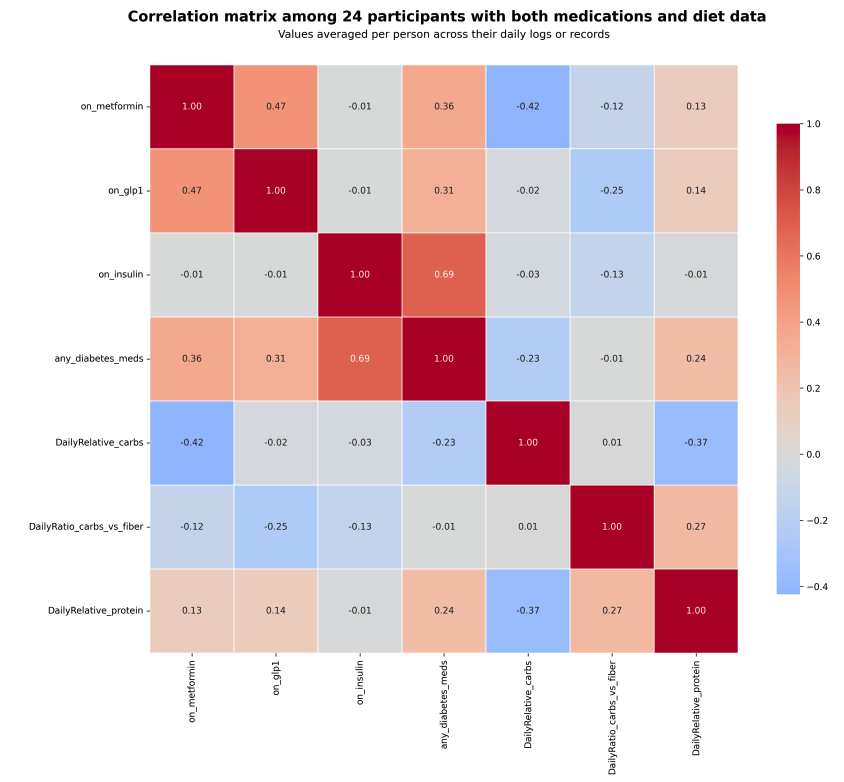

**Supplemental Figure S7.** Spearman correlations for effect modifiers, among 24 participants with both CHR-based medications and self-logged diet data.

**Supplemental Table S1.** Main exposure-outcome association effect estimates from unadjusted, minimally-adjusted, and fully-adjusted GEE models with bootstrap bias-corrected 95% confidence intervals from 1000 bootstrap samples.

| Outcome measures | Exposure groups | β (95% CI), unit varies depending on each outcome variable’s unit | | |
| --- | --- | --- | --- | --- |
|  |  | **Unadjusted model** | **Minimally-adjusted model^a^** | **Fully-adjusted model^b^** |
| Daily mean glucose values, mg/dL | | | | |
| Mean glucose, mg/dL | Regular cycles (ref) | *Ref* | *Ref* | *Ref* |
|  | Irregular cycles | 12.12 (3.19, 20.9) mg/dL | 6.78 (-1.66, 14.29) mg/dL | 6.33 (-1.48, 14.40) mg/dL |
|  | PCOS/PMOS | 8.68 (0.60, 17.29) mg/dL | 7.60 (0.87, 15.48) mg/dL | 6.81 (-0.70, 14.72) mg/dL |
| Daily % glucose observations in, above, or below target glucose range (target range: 70-180 mg/dL) | | | | |
| % Observations in target range | Regular cycles (ref) | *Ref* | *Ref* | *Ref* |
|  | Irregular cycles | -6.18 (-10.63, -1.19) % | -3.14 (-6.94, 1.11) % | -3.08 (-7.11, -0.91) % |
|  | PCOS/PMOS | -4.00 (-8.39, 0.52) % | -3.76 (-7.66, -0.01) % | -3.62 (-8.01, 0.04) % |
| % Observations above target range | Regular cycles (ref) | *Ref* | *Ref* | *Ref* |
|  | Irregular cycles | 6.36 (1.45, 10.80) % | 3.37 (-1.14, 7.18) % | 3.33 (-0.77, 7.41) % |
|  | PCOS/PMOS | 4.42 (-0.09, 8.83) % | 4.00 (0.24, 7.90) % | 3.77 (0.04, 8.14) % |
| % Observations below target range | Regular cycles (ref) | *Ref* | *Ref* | *Ref* |
|  | Irregular cycles | -0.18 (-0.54, 0.18) % | -0.24 (-0.58, 0.11) % | -0.25 (-0.58, 0.11) % |
|  | PCOS/PMOS | -0.42 (-0.76, -0.01) % | -0.24 (-0.58, 0.16) % | -0.15 (-0.50, 0.24) % |
| Measures of glycemic variability from daily glucose standard deviation (SD) or coefficient of variation (%) | | | | |
| Glucose SD, mg/dL | Regular cycles (ref) | *Ref* | *Ref* | *Ref* |
|  | Irregular cycles | 3.96 (-0.06, 7.71) mg/dL | 1.44 (-1.19, 4.28) mg/dL | 1.05 (-1.63, 3.66) mg/dL |
|  | PCOS/PMOS | -0.84 (-3.95, 2.04) mg/dL | 1.31 (-1.25, 3.87) mg/dL | 1.04 (-1.50, 3.49) mg/dL |
| Glucose %CV | Regular cycles (ref) | *Ref* | *Ref* | *Ref* |
|  | Irregular cycles | 1.09 (-0.59, 2.66) % | 0.13 (-0.92, 1.14) % | -0.03 (-1.16, 1.08) % |
|  | PCOS/PMOS | -1.34 (-2.56, -0.08) % | -0.07 (-1.13, 1.03) % | -0.15 (-1.26, 0.87) % |
| Monthly Glucose Management Indicator (GMI) as an approximation of HbA1c (indicating overall glucose control), among 619 participants with monthly GMI for person-months with ≥14 days of CGM data | | | | |
| GMI, % | Regular cycles (ref) | *Ref* | *Ref* | *Ref* |
|  | Irregular cycles | 0.29 (0.07, 0.50) % | 0.16 (-0.02, 0.34) % | 0.15 (-0.01, 0.34) % |
|  | PCOS/PMOS | 0.20 (-0.04, 0.43) % | 0.20 (-0.01, 0.43) % | 0.20 (0.00, 0.42) % |

EL: early-life; CI: confidence interval; GEE, generalized estimation equations; PCOS, polycystic ovary syndrome. In May 2026, PCOS was renamed as polyendocrine metabolic ovarian syndrome (PMOS). We use PCOS throughout this manuscript because it was the term used when survey data were collected, prior to the May 2026 renaming.

^a^ Adjusted for age, self-reported prediabetes, type 1 diabetes, and type 2 diabetes. All covariates are self-reported at enrollment.

^b^ Adjusted for age, self-reported prediabetes, type 1 diabetes, type 2 diabetes, BMI, education level, race/ethnicity, socioeconomic status, typical physical activity level, typical low carb diet preference, and stress reflected by PSS-4 score. All covariates are self-reported at enrollment.

^c^ As GMI is intended to reflect average glycemia over a sufficient CGM wear period and is commonly using at least 14 days of CGM data, we calculated monthly GMI for person-months with ≥14 valid CGM days. Monthly aggregation reduced short-term day-to-day variability while preserving longitudinal information across the monitoring period. GMI estimated HbA1c (%) = 3.31 + 0.02392 × [mean glucose in mg/dL] (Bergenstal et al. 2018. Diabetes Care)

**Supplemental Table S2.** Associations of having PCOS/early-life irregularity (vs. no PCOS and early-life regular) and past or current HbA1c levels, among subsets of participants with linked clinical health records.

| Outcome measures, from clinical health records | Exposure groups | β (95% CI) | | |
| --- | --- | --- | --- | --- |
|  |  | **Unadjusted model** | **Minimally-adjusted model^a^** | **Fully-adjusted model^b^** |
| For past (i.e., pre-CGM) HbA1c levels and variability:  Subset N = 227 participants with clinical HbA1c lab records at any time before their CGM readings (median [IQR]: 3.5 years [1.7–6.2 years] before their CGM readings) | | | | |
| Mean long-term HbA1c, in % of total hemoglobin | Regular cycles (n = 120) | *Ref* | *Ref* | *Ref* |
|  | PCOS/PMOS or irregular cycles (n = 107) | 0.47 (0.03, 0.89) | 0.17 (-0.29, 0.61) | 0.17 (-0.30, 0.66) |
| Among those with ≥3 HbA1c records (N = 162): Long-term HbA1c variability, reflected by coefficient of variation (CV), in % | Regular cycles (n = 83) | *Ref* | *Ref* | *Ref* |
|  | PCOS/PMOS or irregular cycles (n = 79) | 1.8 (-0.6, 4.3) | 0.4 (-2.3, 3.3) | 0.3 (-2.4, 2.7) |
| For concurrent HbA1c levels:  Subset N = 139 participants with clinical HbA1c lab records within +/- 1 month around any of their CGM readings | | | | |
| Mean short-term HbA1c, in % of total hemoglobin | Regular cycles (n = 77) | *Ref* | *Ref* | *Ref* |
|  | PCOS/PMOS or irregular cycles (n = 62) | 0.58 (0.03, 1.22) | 0.35 (-0.18, 0.93) | 0.36 (-0.22, 0.89) |

EL: early-life; CI: confidence interval; PCOS, polycystic ovary syndrome. In May 2026, PCOS was renamed as polyendocrine metabolic ovarian syndrome (PMOS). We use PCOS throughout this manuscript because it was the term used when survey data were collected, prior to the May 2026 renaming.

^a^ adjusted for age, self-reported prediabetes, type 1 diabetes, and type 2 diabetes

^b^ adjusted for age, self-reported prediabetes, type 1 diabetes, type 2 diabetes, BMI, education level, race/ethnicity, SES, physical activity, typical low carb diet preference, and stress reflected by PSS-4 score.

**Supplemental Table S3.** Full covariate adjusted associations between PCOS/irregular cycles and select glycemic outcomes from GEE models with bootstrap bias-corrected 95% confidence intervals from 1000 bootstrap samples, stratified by same-day physical activity levels reflected by Apple Watch activity ring closure status among 521 participants with available same-day wearables data (113,324 days).

| Outcome measures | Exposure groups | β (95% CI) for PCOS/PMOS or EL irregular vs. no PCOS & EL regular, unit varies depending on each outcome variable’s unit^a^ | | | |
| --- | --- | --- | --- | --- | --- |
|  |  | **During person-days with 0 ring closure** | **During person-days with 1 ring closed** | **During person-days with 2 rings closed** | **During person-days with all 3 rings closed** |
| Daily glucose, mg/dL | Regular cycles (ref) | *Ref* | *Ref* | *Ref* | *Ref* |
|  | PCOS/PMOS or irregular cycles | 8.02 (-0.90, 17.36) mg/dL | 4.29 (-2.65, 12.42) mg/dL | 7.61 (-0.86, 17.24) mg/dL | 0.49 (-6.35, 6.23) mg/dL |
| % Observations above target range | Regular cycles (ref) | *Ref* | *Ref* | *Ref* | *Ref* |
|  | PCOS/PMOS or irregular cycles | 4.30 (-1.43, 9.59) % | 2.84 (-1.36, 7.29) % | 3.82 (-0.63, 8.38) % | 0.66 (-3.05, 4.07) % |

EL: early-life; CI: confidence interval; GEE, generalized estimation equations; PCOS, polycystic ovary syndrome. In May 2026, PCOS was renamed as polyendocrine metabolic ovarian syndrome (PMOS). We use PCOS throughout this manuscript because it was the term used when survey data were collected, prior to the May 2026 renaming.

^a^ Adjusted for age, self-reported prediabetes, type 1 diabetes, type 2 diabetes, BMI, education level, race/ethnicity, socioeconomic status, typical physical activity level, typical low carb diet preference, and stress reflected by PSS-4 score. All covariates are self-reported at enrollment.

**Supplemental Table S4.** Covariate-adjusted exposure-outcome association effect estimates, from GEE models with bootstrap bias-corrected 95% confidence intervals from 1000 bootstrap samples, stratified by diabetes status.

| Outcome measures | Exposure groups | β (95% CI) for PCOS/PMOS or EL irregular vs. no PCOS & EL regular, unit varies depending on each outcome variable’s unit^a^ | | |
| --- | --- | --- | --- | --- |
|  |  | **Among those without type 1 or type 2 diabetes** | **Among those with type 1 diabetes** | **Among those with type 2 diabetes** |
| Daily glucose, mg/dL | Regular cycles (ref) | *Ref* | *Ref* | *Ref* |
|  | Irregular cycles | 3.13 (-3.15, 10.24) mg/dL | 4.54 (-5.71, 15.01) mg/dL | 5.77 (-11.18, 27.65) mg/dL |
|  | PCOS/PMOS | 8.02 (-1.23, 21.20) mg/dL | 12.65 (-1.92, 26.62) mg/dL | -2.71 (-15.40, 9.44) mg/dL |
| % Observations above target range | Regular cycles (ref) | *Ref* | *Ref* | *Ref* |
|  | Irregular cycles | 1.43 (-1.38, 5.49) % | 2.82 (-2.64, 8.41) % | 2.08 (-7.02, 13.98) % |
|  | PCOS/PMOS | 3.79 (0.07, 9.93) % | 6.34 (-0.39, 12.71) % | -0.73 (-8.28, 6.92) % |

CI: confidence interval; GEE, generalized estimation equations; PCOS, polycystic ovary syndrome. In May 2026, PCOS was renamed as polyendocrine metabolic ovarian syndrome (PMOS). We use PCOS throughout this manuscript because it was the term used when survey data were collected, prior to the May 2026 renaming.

^a^ Adjusted for age, self-reported prediabetes, type 1 diabetes, type 2 diabetes, BMI, education level, race/ethnicity, socioeconomic status, typical physical activity level, typical low carb diet preference, and stress reflected by PSS-4 score. All covariates are self-reported at enrollment.

**Supplemental Table S5.** Full covariate adjusted associations further evaluating PCOS with or without androgen excess symptoms, from GEE models with robust sandwich 95% CIs.

| Outcome measures | Exposure groups | No. among the 661 participants | β (95% CI), unit varies depending on each outcome variable’s unit^a^ |
| --- | --- | --- | --- |
| Daily mean glucose values, mg/dL | | | |
| Mean glucose, mg/dL | Regular cycles (ref) | 412 (ref) | *Ref* |
|  | Irregular cycles | 137 | 6.37 (4.31, 8.43) mg/dL |
|  | PCOS/PMOS, without AE symptoms | 25 | 4.33 (1.01, 7.66) mg/dL |
|  | PCOS/PMOS, with AE symptoms^b^ | 87 | 7.57 (5.33, 9.80) mg/dL |
| Daily % observations in, above, or below target glucose range (target range: 70-180 mg/dL) | | | |
| % Observations in target range | Regular cycles (ref) | 412 (ref) | *Ref* |
|  | Irregular cycles | 137 | -3.09 (-4.09, -2.09) % |
|  | PCOS/PMOS, without AE symptoms | 25 | -2.57 (-4.41, -0.73) % |
|  | PCOS/PMOS, with AE symptoms^b^ | 87 | -3.95 (-5.10, -2.79) % |
| % Observations above target range | Regular cycles (ref) | 412 (ref) | *Ref* |
|  | Irregular cycles | 137 | 3.34 (2.34, 4.35) % |
|  | PCOS/PMOS, without AE symptoms | 25 | 2.78 (0.94, 4.63) % |
|  | PCOS/PMOS, with AE symptoms^b^ | 87 | 4.08 (2.93, 5.24) % |
| % Observations below target range | Regular cycles (ref) | 412 (ref) | *Ref* |
|  | Irregular cycles | 137 | -0.25 (-0.38, 0.12) % |
|  | PCOS/PMOS, without AE symptoms | 25 | -0.20 (-0.44, 0.05) % |
|  | PCOS/PMOS, with AE symptoms^b^ | 87 | -0.14 (-0.32, 0.04) % |
| Measures of glycemic variability | | | |
| Glucose SD, mg/dL | Regular cycles (ref) | 412 (ref) | *Ref* |
|  | Irregular cycles | 137 | 1.02 (0.40, 1.63) mg/dL |
|  | PCOS/PMOS, without AE symptoms | 25 | 3.46 (1.86, 5.06) mg/dL |
|  | PCOS/PMOS, with AE symptoms^b^ | 87 | 0.30 (-0.34, 0.94) mg/dL |
| Glucose %CV | Regular cycles (ref) | 412 (ref) | *Ref* |
|  | Irregular cycles | 137 | -0.05 (-0.34, 0.24) % |
|  | PCOS/PMOS, without AE symptoms | 25 | 1.35 (-0.76, 1.95) % |
|  | PCOS/PMOS, with AE symptoms^b^ | 87 | -0.61 (-1.90,-0.32) % |

EL: early-life; CI: confidence interval; GEE, generalized estimation equations; AE, androgen excess; PCOS, polycystic ovary syndrome. In May 2026, PCOS was renamed as polyendocrine metabolic ovarian syndrome (PMOS). We use PCOS throughout this manuscript because it was the term used when survey data were collected, prior to the May 2026 renaming.

We imputed AE status for 55 participants with PCOS/PMOS who did not provide information relevant to whether or not they had potential androgen excess symptoms, using multiple imputations by chained equations (50 imputed sets). Because combining multiple imputation with bootstrapping was computationally intensive, secondary analyses stratified by AE status used cluster-weighted generalized estimating equation models with robust sandwich 95% CIs (i.e., without bootstrapping). Accordingly, results should be interpreted primarily based on the magnitude and direction of the point estimates. Estimates from the 50 imputed datasets were pooled according to Rubin’s rules.

^a^ Adjusted for age, self-reported prediabetes, type 1 diabetes, type 2 diabetes, BMI, education level, race/ethnicity, socioeconomic status, typical physical activity level, typical low carb diet preference, and stress reflected by PSS-4 score. All covariates are self-reported at enrollment.

^b^ Presence of androgen excess symptoms include any self-reported potential signs of hirsutism (reported hair growth on the body; or reported several or a lot of hair growth on upper lip or the chin) or moderate/severe acne (reported red, irritated pimples; pimples with pus; or scars from acne) in the survey questions relevant to self-evaluated hormonal symptoms.

**Supplemental Table S6.** Summary of data sources of main exposure, outcome, covariate, and effect modifiers.

| Variable category in analyses | Variables | Data type and source | Data source, details | Additional notes |
| --- | --- | --- | --- | --- |
| Main outcomes | Daily: mean glucose levels, %observations in, above, or below target range, glucose SD, glucose %CV;  Monthly: glucose management indicator (GMI) | Wearables  (Apple Health) | **Continuous glucose monitoring (CGM)** device data  (99.6% from Dexcom G6; 0.4% from FreeStyle Libre) | Monthly GMI derived from CGM readings in months with ≥14 days of CGM data |
| Main exposures | 3-Level mutually exclusive groups:^a^  1) regular cycles; 2) irregular cycles; 3) PCOS/PMOS | Survey | Baseline Medical History Survey  Baseline Reproductive History Survey | N/A |
| Secondary exposures | PCOS group further stratified by androgen excess symptoms (yes/no) | Survey | Baseline Hormonal Symptoms Survey | Only subsets with available data used |
| Covariates | Age, self-reported prediabetes, type 1 diabetes, type 2 diabetes, BMI, education level, race/ethnicity, socioeconomic status, typical physical activity level, typical low carb diet preference, and stress reflected by PSS-4 score | Survey | Baseline Demographics Survey  Baseline Medical History Survey  Baseline Health Overview Survey | All covariates are based on self-report at study enrollment |
| Proposed effect modifiers | Same-day physical activity:  0, 1, 2, or 3 daily activity ring closure | Wearables/Sensor  (Apple Watch) | **Apple Watch activity ring closure** data:  Move; Exercise; Stand | Only subsets with available data used |
|  | Same-day diet:  Daily macronutrient composition | Digital log  (Apple Health) | **Dietary log** data shared in the **Apple Health app**  (Varying sources: Apple Health app, and any participating third-party dietary/wellness tracking app that is permitted to be shared in the Apple Health app) | Only subsets with available data used |
|  | Medication use during CGM monitoring:  Metformin, GLP-1 receptor agonists, insulin, other common diabetes medications | Electronic health records  (Apple Health) | **Medication record** data from linked **clinical health records (CHR)** that participants shared with the study | Only subsets with available data used; as CHRs are voluntarily shared, data included in this analyses may or may not include a person’s comprehensive CHR over the life course. |
| Other relevant cardiometabolic vital signs | HbA1c levels in blood, +/- 1 month around CGM monitoring | Electronic health records  (Apple Health) | **Laboratory test** data from linked **clinical health records (CHR)** that participants shared with the study | Only subsets with available data used; as CHRs are voluntarily shared, data included in this analyses may or may not include a person’s comprehensive CHR over the life course. |
|  | Same-day resting heart rate levels | Wearables/Sensor  (Apple Watch) | **Wearables/sensor:**  **Apple Watch activity ring closure** data | Only subsets with available data used |

PCOS, polycystic ovary syndrome. In May 2026, PCOS was renamed as polyendocrine metabolic ovarian syndrome (PMOS). We use PCOS throughout this manuscript because it was the term used when survey data were collected, prior to the May 2026 renaming.

^a^ Regular cycles group: those without self-reported PCOS diagnosis, and reported spontaneously establishing regular cycles within 4 years after menarche; Irregular cycles group: those without self-reported PCOS diagnosis, and reported reaching cycle regularity in 5+ years after menarche, their cycles are still irregular, or used hormones to achieve regularity.
